# Distribution and transmission of *M. tuberculosis* in a high-HIV prevalence city in Malawi: a genomic and spatial analysis

**DOI:** 10.1101/2024.05.17.24307525

**Authors:** Melanie H. Chitwood, Elizabeth L. Corbett, Victor Ndhlovu, Benjamin Sobkowiak, Caroline Colijn, Jason R. Andrews, Rachael M. Burke, Patrick G.T. Cudahy, Peter J. Dodd, Jeffrey W. Imai-Eaton, David M. Engelthaler, Megan Folkerts, Helena Feasey, Yu Lan, Jen Lewis, Nicolas A Menzies, Geoffrey Chipungu, Marriott Nliwasa, Daniel M. Weinberger, Joshua L. Warren, Joshua A. Salomon, Peter MacPherson, Ted Cohen

## Abstract

**Background:** Delays in identifying and treating individuals with infectious tuberculosis (TB) contribute to poor health outcomes and allow ongoing community transmission of *M. tuberculosis* (*Mtb*). Current recommendations for screening for tuberculosis specify community characteristics (e.g., areas with high local tuberculosis prevalence) that can be used to target screening within the general population. However, areas of higher tuberculosis burden are not necessarily areas with higher rates of transmission. We investigated the genomic diversity and transmission of *Mtb* using high-resolution surveillance data in Blantyre, Malawi.

**Methods and Findings:** We extracted and performed whole genome sequencing on mycobacterial DNA from cultured *M. tuberculosis* isolates obtained from culture-positive tuberculosis cases at the time of tuberculosis (TB) notification in Blantyre, Malawi between 2015-2019. We constructed putative transmission networks identified using TransPhylo and investigated individual and pair-wise demographic, clinical, and spatial factors associated with person-to-person transmission. We found that 56% of individuals with sequenced isolates had a probable direct transmission link to at least one other individual in the study. We identified thirteen putative transmission networks that included five or more individuals. Five of these networks had a single spatial focus of transmission in the city, and each focus centered in a distinct neighborhood in the city. We also found that approximately two-thirds of inferred transmission links occurred between individuals residing in different geographic zones of the city.

**Conclusion:** While the majority of detected tuberculosis transmission events in Blantyre occurred between people living in different zones, there was evidence of distinct geographical concentration for five transmission networks. These findings suggest that targeted interventions in areas with evidence of localized transmission may be an effective local tactic, but will likely need to be augmented by city-wide interventions to improve case finding and to address social determinants of tuberculosis to have sustained impact.

**Author Summary:** *Why was this study done?*

– Tuberculosis (TB) is a major global health threat and a leading cause of death due to infectious disease. Rapid diagnosis and treatment of individuals with TB is vital to reduce the spread of disease.
– If public health programs can identify areas with ongoing TB transmission, resources might be directed toward intervening in those areas to interrupt transmission chains. However, in settings where many people have TB, it is often difficult to differentiate areas with high rates of disease from areas with high rates of local transmission.

*What did the researchers do and find?*

– We used whole genome sequencing data to infer networks of TB transmission in Blantyre, Malawi. We used individual residence data to identify whether transmission networks were concentrated in specific parts of the city and to describe the amount of transmission that occurred between vs. within distinct parts of the city.
– We found that most TB transmission in Blantyre occurred between individuals who did not live near each other. We also identified five transmission networks which had strong local foci of transmission.

*What do these findings mean?*

– Because most TB transmission in Blantyre does not occur in concentrated areas, city-wide interventions, such as improving access to TB care services and addressing social determinants of TB, may be needed to improve TB control.
– For areas where there is evidence of local concentrated transmission, additional resources and strategies, such as targeted active case finding, may help to more rapidly reduce transmission and TB incidence.

## Introduction

Tuberculosis (TB) is a major global health threat and a leading infectious cause of death. The World Health Organization’s (WHO) End TB Strategy aims to reduce global tuberculosis incidence by 80% by 2030 from 2015 levels^(1)^. Rapid diagnosis and treatment, key pillars of the End TB Strategy, can reduce tuberculosis transmission by limiting the time an individual is infectious and potentially transmitting *M. tuberculosis* (*Mtb*). Passive case detection, which depends on individuals with tuberculosis seeking care, is insufficient to rapidly reduce *Mtb* transmission in most settings^(2)^. In high-burden settings, WHO recommends systematic screening for tuberculosis disease in communities^(3)^. However, there is inconsistent evidence to indicate whether screening decreases tuberculosis prevalence^(4, 5)^.

Targeting screening in areas where most transmission occurs may decrease TB prevalence^(6, 7)^, but identifying these areas using routinely collected data is challenging. Among newly infected individuals, the incubation period is variable and the risk of progressing to symptomatic disease is generally low^(8)^. Areas of high disease burden may therefore reflect higher risk of progression to active disease rather than higher risk of transmission^(9)^. This phenomenon may be more pronounced in high human immunodeficiency virus (HIV) prevalence settings because people living with HIV are more likely to progress to active tuberculosis disease^(10)^ and less likely to transmit *Mtb* to others^(11)^. Consequently, methods are needed that can identify areas of active transmission, which may not necessarily align with areas of high notification rates.

The increasing availability of whole genome sequencing (WGS) data, paired with methodological advances in transmission inference, has improved the ability to understand pathogen transmission dynamics^(8, 12, 13)^, information that is critical for the design of targeted active case finding efforts. Several studies have leveraged these types of data to characterize spatial patterns in tuberculosis burden and *Mtb* transmission^(14–19)^, but few have been conducted in cities with high rates of tuberculosis/HIV co-infection^(20)^.

In this study, we collected and sequenced mycobacterial specimens from individuals diagnosed with culture-positive tuberculosis in Blantyre, Malawi between 2015 and 2019. We used WGS data to infer networks of transmission and paired those findings with geographical coordinates (GPS) of patient home locations to identify local transmission of specific strains and describe patterns of transmission between administrative areas. We hypothesized that whole genome sequencing would provide novel insights into *Mtb* transmission dynamics in a city with a high prevalence of both tuberculosis and HIV.

## Methods

### Study setting and population

Blantyre is a city in southern Malawi, with a population of approximately 800,000^(21)^. It is the second largest city in Malawi and is the nation’s industrial and commercial capital. Over half of the population live in areas without access to basic municipal services and 43% of city land is considered unplanned or rural^(22)^. Blantyre is a hilly city, and its varied topography creates distinct neighborhoods separated by ridges and valleys.

The study protocols were reviewed and approved by the University of Malawi College of Medicine Research and Ethics Committee (#P.12/18/2556), the London School of Hygiene and Tropical Medicine (#16228-4), and Yale (#2000028431). Oral consent was provided by people registering for TB treatment for electronic data capture, including recording of household co-ordinates. Oral consent and assent were used for the latter since the electronic register data capture was conducted as part of normal clinical practice by District TB Officers. Approval was sought and gained from the District Health Office and local chiefs for the study to be conducted in their areas.

This study included people diagnosed with tuberculosis in Blantyre, Malawi between 1 January 2015 and 31 December 2019. All people with notified tuberculosis in Blantyre were registered in the ePAL (electronic Participant Locator) system^(23–25)^. ePAL is an app-based data entry platform for the collection of patient information combined with an electronic case report form with high resolution satellite maps and community-identified points of interest^(25)^. Data available in ePAL include age, sex, diagnosing clinic, microbiology results (acid-fast bacillus [(AFB]) smear and Xpert MTB/RIF), symptom history and duration, tuberculosis classification (pulmonary, extra-pulmonary), HIV and antiretroviral therapy (ART) status, HIV clinic (if applicable), presence of known TB exposures (e.g. household contacts), locations of the three most recent clinics attended, number of hospital admissions within the year preceding diagnosis, and a range of poverty indicators. The patient’s home location, selected via touch-screen and converted in-app to GPS coordinates^(25)^, is also available.

We estimated tuberculosis notification rates at a resolution of 500m^2^ grid cells. Population denominators were calculated from WorldPop^(21)^ 2020 data and aggregated from the original resolution of 100m^2^. We also present grid-specific HIV prevalence estimates, which were derived from two national HIV prevalence studies, one Blantyre-specific prevalence study, and antenatal prevalence data. These data were combined in a Bayesian model to derived highly spatially resolved HIV prevalence estimates, as described previously^(26)^.

### Laboratory regrowth, DNA isolation, and whole genome sequencing

We set out to re-culture and sequence samples from all culture-positive cases notified over the five-year study period. Study isolates were thawed from −80°C and cultured in liquid Middlebrook 7H9 media using the BD BACTECT™ MGIT™ 960 system, then subcultured on Löwenstein–Jensen medium at 37°C to obtain pure colonies. A minimum of 1 µg *Mtb DNA* (either 100µl of 10000 ng/ml, or 40µl of 25000 ng/ml, etc) was manually extracted following standard operating procedures as previously described^(27)^ and validated in-country, with DNA stored at −20°C before shipping for sequencing at TGen in Flagstaff, Arizona.

Sequencing libraries were be constructed using either Illumina’s DNA Prep kit or Watchmaker’s DNA Library Prep Kit with Fragmentation. Whole genome sequencing (WGS) was performed on an Illumina NextSeq550 or NextSeq1000 to produce paired-end, 150bp reads. A phiX (Illumina) sequencing control was spiked into each run at 1% of the total library to be sequenced to facilitate run performance monitoring. Raw sequencing FASTQ files were checked for non-Mycobacterium DNA using Kraken;^(28)^ isolates containing > 80% *Mtb* reads were retained and non-*Mtb* reads filtered out using a custom script. Sequence reads were mapped to the H37Rv reference strain (GenBank accession number NC_000962.3) with BWA v.0.7.17 ‘mem’^(29)^, removing sequences with < 80% mapping to the reference strain and < 50x average coverage.

Variant calling was carried out using GATK v.4.4.0.0 ‘HaplotypeCaller’ and ‘GenetypeGVCFs’^(30)^. Single nucleotide polymorphisms (SNPs) were filtered to remove sites with low quality (Q < 20), low read depth (DP < 5), or high proportion missingness (missing call in ≥ 10% of isolates). Sites showing more than one allele (mixed sites) were assigned the majority allele where ≥ 90% of reads agreed, otherwise these were assigned an ambiguous character ‘N’. Finally, sequences with a high likelihood of mixed infection identified using MixInfect^(31)^ were removed. *In silico* lineage prediction and drug resistance profiling was carried out on the remaining isolates using TB-Profiler v.5.0.1^(32)^.

### Phylogenetic reconstruction

A multi-sequence alignment of concatenated SNPs was used to produce phylogenetic trees. SNPs in repetitive regions and known microbial resistance-associated and PE/PPE genes were removed to account for potential homoplasy that may confound phylogenetic reconstruction (Supplemental Table 1). IQ-tree v.2.2.2.6^(33)^ was used to construct a maximum-likelihood phylogeny of all isolates, with the ‘-m TEST’ parameter used to determine the optimal nucleotide substitution model of Kimura’s model with unequal base frequencies (K3Pu+F).

### Transmission inference

Transmission networks and the probability of person-to-person transmission among sequenced cases was inferred using a two-step process. First, we identified preliminary, broad clusters of sequences using a 50 SNP threshold and constructed timed phylogenies with BEAST2 v.2.7.5^(34)^. We used a relaxed lognormal substitution rate and ran the model for 2×10^8^ Markov chain Monte Carlo (MCMC) iterations or until convergence was achieved and an adequate number of posterior samples were collected, demonstrated by the collected samples from all parameters reaching an effective sample size ESS of greater than 200 after a 20% burn-in was discarded.

Second, we ran TransPhylo^(35)^ on each broad cluster to identify transmission networks. We used the implementation of TransPhylo with simultaneous inference of multiple trees ^(14)^ to account for phylogenetic uncertainty by taking a random sample of 50 posterior trees from the BEAST2 output, discarding the first 20% as burn-in. We assumed a prior gamma generation time distribution (α =1.3, β = 0.3) and a prior gamma sampling time distribution (α =1.1, β = 0.4), as has been previously applied for *Mtb* transmission reconstruction^(35–37)^. We ran the model for 1×10^5^ MCMC iterations using a fixed within-host coalescent parameter of 100/365 and a beta sampling proportion distribution (α = 2, β = 20) that updated through the runs. This produced a predicted pairwise probability of direct transmission between isolates in clusters, and all isolates that were not present in the same broad cluster were *de facto* assigned a pairwise transmission probability of 0.

Transmission networks predicted by TransPhylo for each broad cluster with the highest probability were further refined. Where inferred networks were linked by more than three non-sampled hosts, we considered these as separate networks. This allowed us to identify putative transmission networks while accounting for missing cases.

### Identification of spatial foci of transmission

For each putative transmission network with at least five cases, we investigated spatial areas where individuals with tuberculosis had relatively higher likelihoods of transmission to or from others in the area. We call these areas spatial foci of transmission, and we identify them using a non-parametric distance-based mapping (DBM)^(38)^ approach implemented in the R package hotspotr^(39)^. Using hotspotr, we divided the city into a 100 by 100 grid of cells (each cell is 194 m by 144 m). The analysis then proceeds for each transmission network. First, we select a transmission network to analyze. Second, we use the home location of tuberculosis cases that were not part of that transmission network to calculate the expected number of cases within each grid cell. Third, we calculate the risk that there are more cases belonging to the transmission network in a given grid cell than expected, assigning a score between 0 (no spatial aggregation of individuals from the same transmission network) and 1 (highest risk of spatial aggregation). We repeated these steps for each transmission network with five or more individuals. Any grid cell with a score ≥ 0.95 was considered a spatial focus of transmission, and we used a narrow window (width = 0.01) to smooth results across grid cells^(39)^. Note that this method allows that there may be multiple foci of transmission for a single transmission network.

### Pairwise regression analysis of transmission networks

We used the R package GenePair^(40)^ to quantify the association between the genetic relatedness of pairs of cases and several pair- and individual-level factors, including spatial proximity and shared healthcare clinics. Regression models with dyadic (paired) outcomes will produce overly optimistic effect estimates if the correlation between dyadic outcomes is not properly accounted for^(40, 41)^; GenePair solves this problem by including spatially-structured individual-level random effect parameters within a regression framework that induce correlation between the dependent variables^(40)^.

We fit a single regression model with every individual in a transmission network, using a binary outcome indicator of whether two individuals were in the same network. We included predictors (age, sex, HIV status, HIV clinic, tuberculosis clinic, home address) if there were no missing values of that predictor among individuals in the transmission network, and if there were more than four pairs of individuals in each level of the predictor. We allowed the impact proximity on the relationship between cases to differ for cases that were close together (< 6km) and cases that were further apart (≥ 6km). We chose to model distance in this way because we believe the association of distance and cluster membership may be stronger at shorter distances. We performed model comparisons using AIC, and found that a 6km threshold outperformed a single linear effect and also other choices of threshold.

In a separate analysis, we fitted regression models for each transmission network with at least 10 cases, using the number of SNPs by which pairs differ as the outcome. This allows us to investigate the risk of specific predictors on SNP distance conditional on belonging to the same transmission network. We use the same set of covariates as above, and also include the zone in which the individuals resides. For all analyses, we based inference on samples from the joint posterior distribution, removing 10,000 iterations of burn-in and thinning the remaining 25,000 by a factor of 5 to reduce correlation in the posterior samples.

### Transmission flow analysis

We divided the city into seven zones based on existing administrative boundaries and expert opinion about intracity mobility (Supplemental Figure 1). We used the R package PhyloFlows^(42)^ to estimate transmission within and between these zones, accounting for the population size and sequencing coverage of each zone (Supplemental Table 2). Zone-level population sizes were calculated by aggregating across grid cells (see ‘Study setting and population’). We fitted PhyloFlows using a transmission probability threshold of 10% and assuming only one infector per individual. We ran the model for 20,000 MCMC iterations.

## Results

Between January 2015 and December 2019, 11,137 tuberculosis disease episodes among individuals aged ≥15 were notified and recorded in ePAL. The mean tuberculosis notification rate over the study period was 279 per 100,000 population per year, which declined from 299 per 100,000 in 2015 to 224 per 100,000 in 2019. The notification rate varied across city zones, and grid cells with high notification rates abutted areas of apparent lower burden (Figure 1A). Among individuals notified in ePAL, most were new cases (12,046; 87%), most were male (7,438; 61%), the median age was 35 years (IQR: 26, 43), and most were living with HIV (7,860; 64%). Areas of higher HIV prevalence in the general populations in Zone 2 and Zone 6 overlapped with areas with high tuberculosis notification rates (Figure 1B). Population coverage of ART is estimated to be greater than 50%^(43)^.

**Figure 1.**
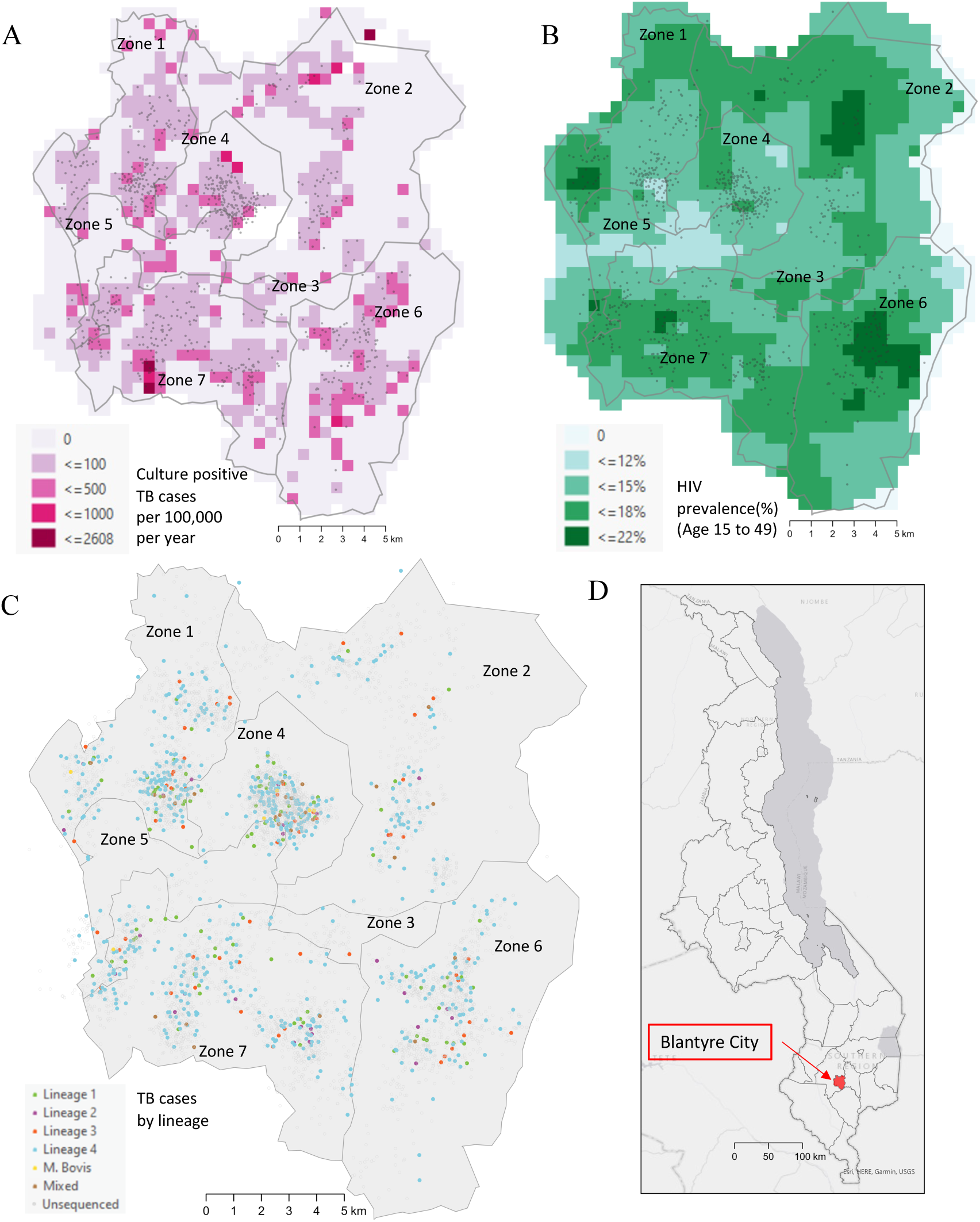
Map of Blantyre, Malawi with **(A)** culture positive tuberculosis case notifications per 100,000 population per year over the study period; **(B)** HIV prevalence estimates (population aged 15-49 years) by zone in the urban area of Blantyre; **(C)** all notified tuberculosis cases, colored by major lineage or shown in grey if no sequencing data were available; **(D)** inset map indicating the location of Blantyre in Malawi. Points have been jittered for privacy.

### Whole genome sequencing analysis

Over the study period, 3,856 individuals tested positive by tuberculosis culture, 1,333 (34.6%) specimens were available for whole genome sequencing, and 1,009 (26.2%) samples could be matched to patient clinical data (Supplemental Figure 2). Among sequenced isolates, 861 (85%) passed quality control checks and 717 (83%) were found to be pure (non-mixed) samples, which were included in the final sample dataset. Home location data were available for 707 (99%) of these cases (Figure 1C).

A freezer failure, during which alarms were not acted on (due to COVID-19 lockdowns) occurred in 2020, resulting in lower-than-expected sequencing yield for affected frozen *Mtb* isolates. The fraction of individuals with a successfully sequenced diagnostic specimen did not differ meaningfully by sex, age, HIV status (Supplemental Table 3A), or city zone of residence (Supplemental Table 3B). However, the fraction of successfully sequenced culture positive cases varied by year, ranging from 12% to 33% (Supplemental Table 3C).

Most isolates included in the final sample dataset belonged to lineage 4 (Euro-American, 72%) followed by lineage 1 (Indo-Oceanic, 14%) (Figure 2), similar to previously reported studies in Blantyre^(44)^. The proportion of sequences collected from each lineage remained consistent throughout the study period (Supplemental Figure 3). There was a low prevalence of drug resistance, with 94% (676/717) of samples susceptible to all antimicrobials (Figure 2). We identified 2 rifampin mono-resistant, 22 isoniazid mono-resistant strains, and 4 multidrug resistant strains (< 1%).

**Figure 2.**
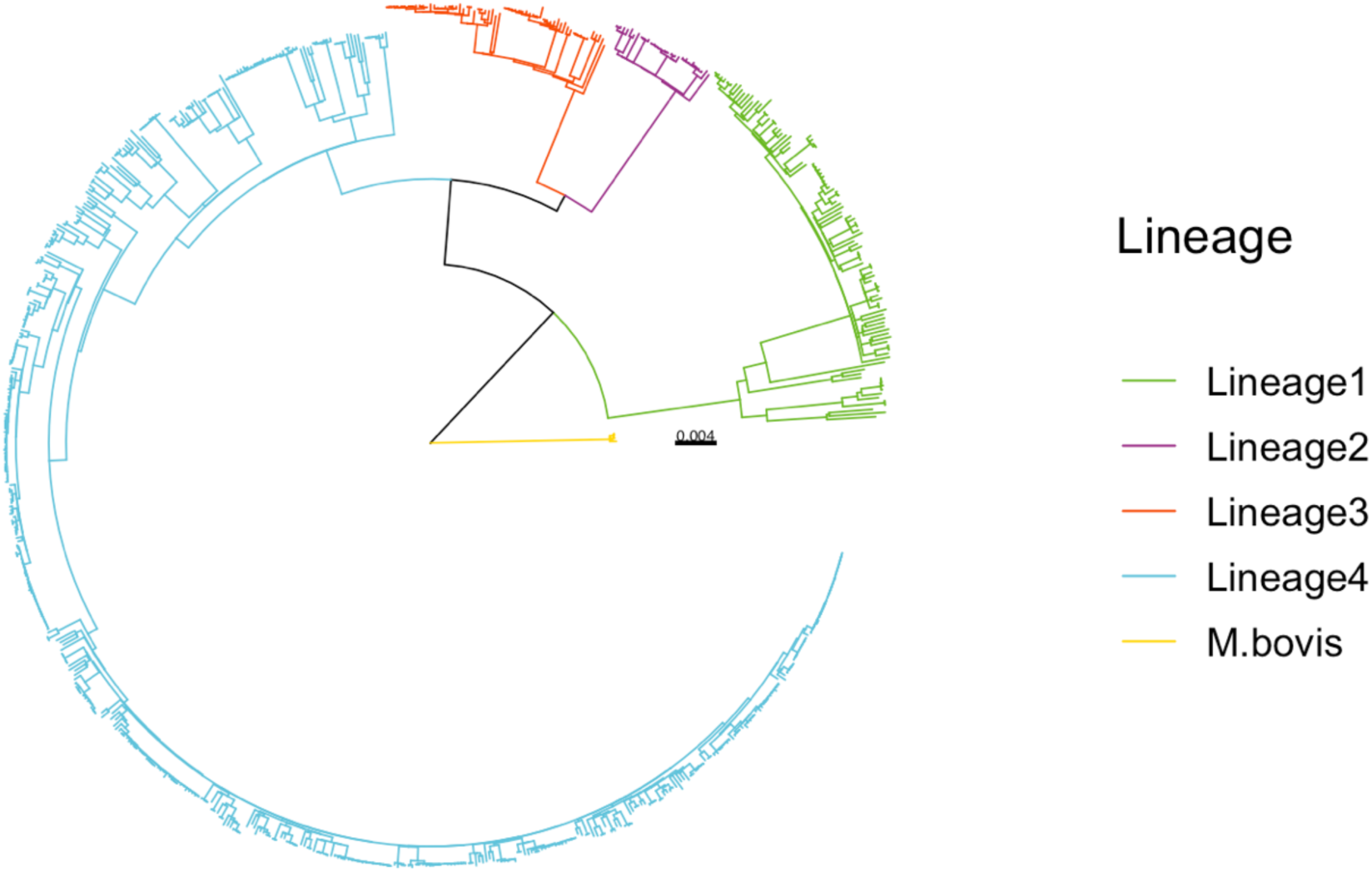
A maximum likelihood phylogeny illustrating the genetic relatedness of the 717 isolates included in the study, colored by major *Mycobacterium tuberculosis* complex lineage. Branch lengths are scaled by substitutions per site.

### Transmission inference

There were 393 isolates (56%) that belonged to one of 130 transmission networks inferred with TransPhylo and had an associated GPS coordinate. Most transmission networks contained only one pair of individuals (87/130; 67%); 13 transmission networks comprised five or more individuals (10%) and three transmission networks comprised 10 more individuals (2%) (Figure 3). The largest transmission network contained 25 individuals. There were 50 pairs of individuals with a high probability of direct transmission (probability ≥ 0.5) and a further 97 pairs with a moderate probability of direct transmission (probability ≥ 0.25).

**Figure 3.**
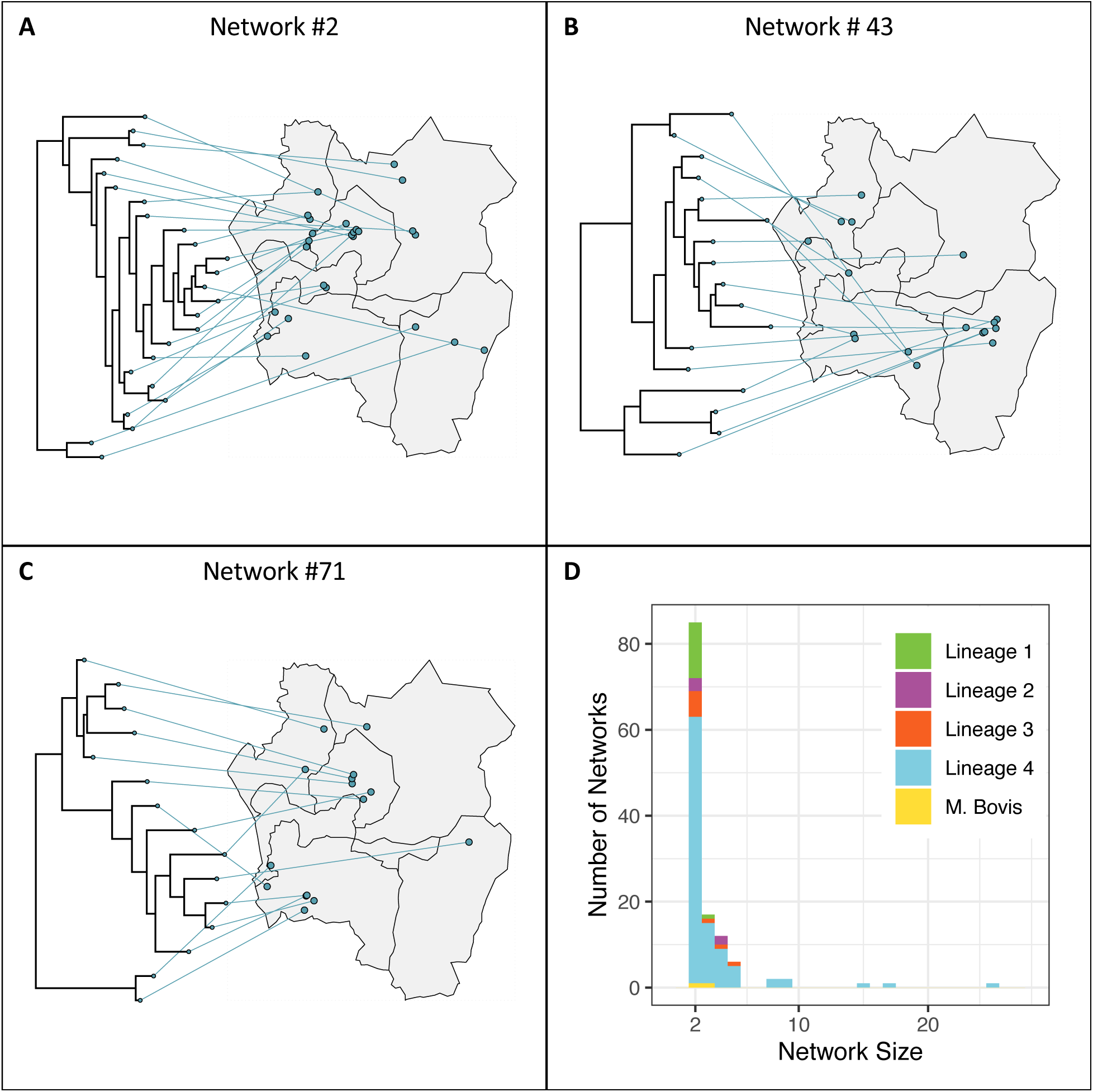
**(A-C)** Time-resolved phylogenies with taxa linked to case home location, colored by main lineage. Each panel contains a different transmission network. The three networks with 10 or more individuals are shown. Points have been jittered for privacy. **(D)** Distribution of transmission network sizes (number of sampled hosts within the network), colored by main lineage.

### Identification of spatial foci of transmission

In distance-based mapping (DBM) of 13 transmission networks with ≥ 5 individuals (Supplemental Table 4), we detected spatial foci of recent transmission in five transmission networks (Figure 4). For the largest transmission network (network 2, n = 25), we did not detect any spatial aggregation of cases, suggesting that it was widespread throughout the city.

**Figure 4.**
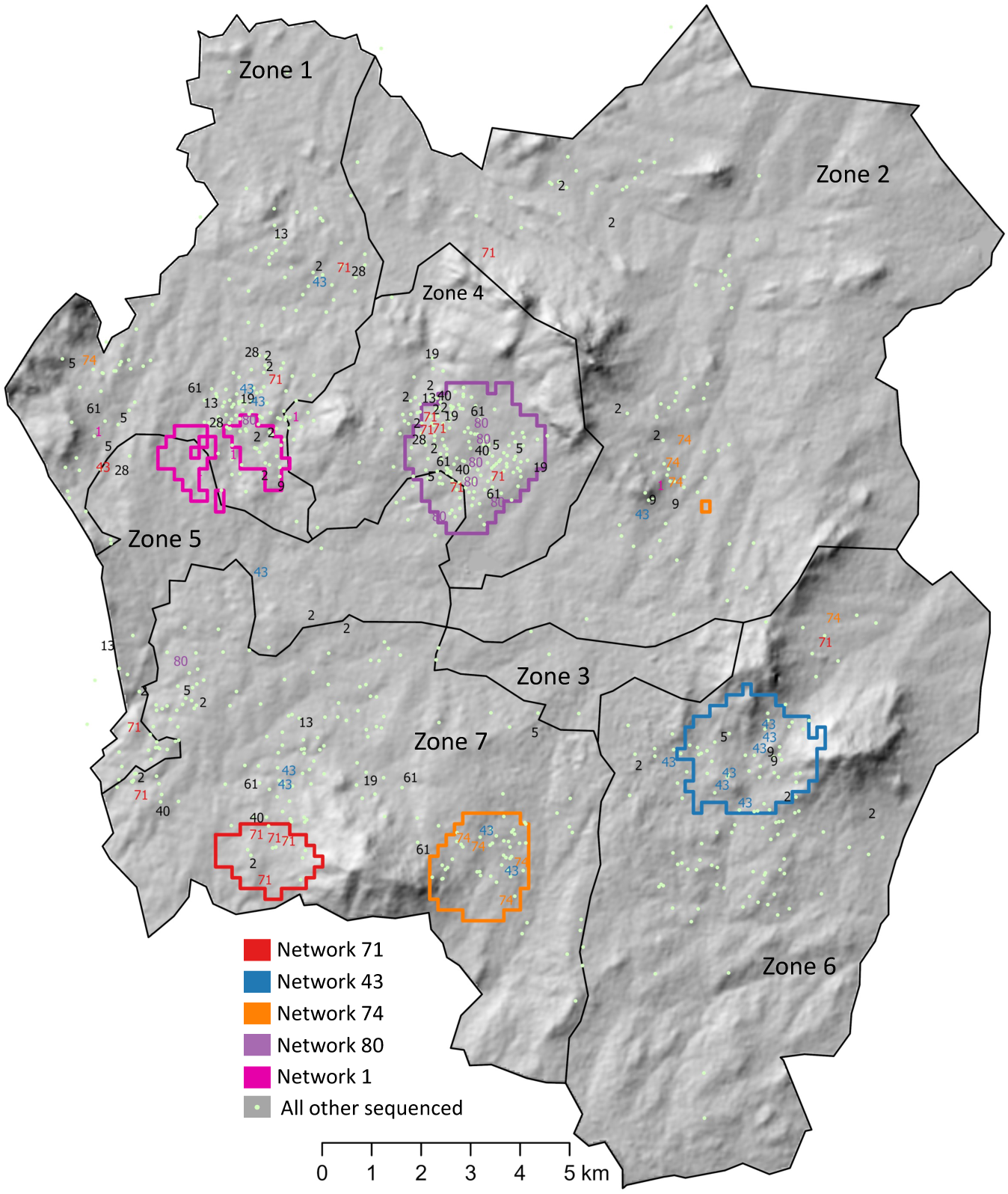
Foci of recent transmission identified using a distance based mapping (DBM) approach based on home address, applied to transmission networks with 5 or more individuals. Circled regions represent areas where the risk of spatial aggregation for a network is greater than 95% (spatial foci). A single transmission network may have multiple spatial foci. Individuals belonging to a transmission network with a spatial focus may reside outside the focus. Points have been jittered for privacy. Base map citation: “Malawi SRTM DEM 30meters.” n.d. Accessed April 24, 2024. https://www.africageoportal.com/datasets/rcmrd::malawi-srtm-dem-30meters/about.

Transmission network #47 was the largest network (n = 17) for which we identified a spatial focus. Most individuals in this network were people living with HIV (n = 10, 67%) and the single spatial focus of transmission was in a high HIV prevalence area. Network #71 was the second largest transmission network, and had a spatial focus on the periphery of the city where there was a high case notification rate. Only 20% (n = 3) of individuals within network #71 were people living with HIV.

Not all members of a transmission cluster with a spatial focus of transmission lived in or near the focus we identified (Supplemental Table 4). In network #80, 75% of network members (n = 6) lived inside the focus, and, in network #1, 60% of network members (n = 3) lived within 1km of the focus boundary (Figure 3A). Overall, of the 393 individuals with TB belonging to a transmission network, 252 (64%) lived within 1 km of a transmission focus.

### Pairwise regression analysis of transmission networks

We analyzed all individuals belonging to a putative transmission network (excluding individuals with missing home location data; n = 389), and found that two individuals had a higher odds of belonging to the same putative transmission network if they shared a tuberculosis diagnostic clinic (adjust odds ratio (aOR) 1.61 95% highest posterior density interval (HPD) 1.14, 2.13), were both HIV negative (aOR 1.51, 95% HPD 1.09, 2.02), and were both male (aOR 1.45, 95% HPD 1.09, 1.91) (Figure 5A). A 1 km increase in the distance between home location was associated with a 0.84 (0.79, 0.88) aOR of belonging to the same cluster for distances up to 6km. Each 1 km increase in distance above the 6 km threshold was associated with a 0.91 (0.89, 0.94) aOR. We did not find a significant association between shared HIV clinic and belonging to the same putative transmission cluster (aOR 0.86, 95% HPD 0.58, 1.21). Because there was very high coverage of antiretroviral therapy among HIV positive individuals in this study (91%), we did not include ART as a covariate in our model.

**Figure 5.**
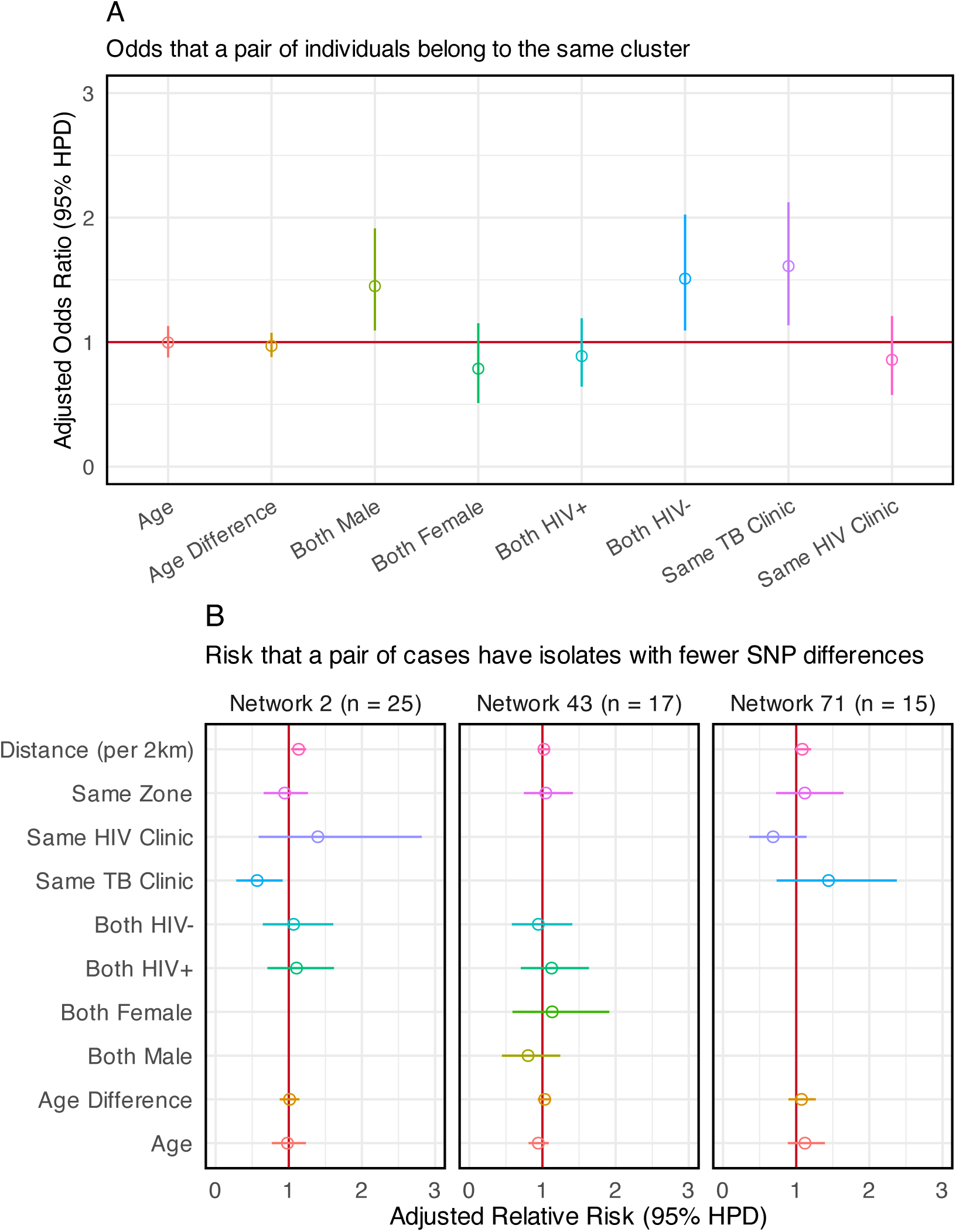
**(A)** Effect of covariates on the odds that case pairs belong to the same putative transmission cluster. Adjusted Odds Ratio > 1 is associated with increased odds of belonging to the same putative transmission cluster. **(B)** Effect estimates of covariates on SNP difference in isolates from case pairs within the same transmission network. Adjusted Relative Risk < 1 is associated with smaller SNP differences on average; smaller SNP differences are associated with increased likelihood of direct transmission between case pairs. Gender is excluded in the analyses of network 2 and network 71 because only three individuals were female. HIV status is excluded in the analysis of network 71 because only three individuals were people living with HIV.

We also analyzed the three largest transmission networks (#2, #43, and #71) in separate analyses (Figure 5B). In network #2, increasing distance between participant homes was associated with a small but significant increase in the number of SNPs by which the sequences differed (adjusted Relative Risk (aRR) per 2 km increase of 1.13, 95% HPD 1.03, 1.23). In addition, individuals in transmission network #2 who were diagnosed at the same tuberculosis clinic had sequences with smaller SNP differences on average (aRR 0.57, 95% HPD 0.28, 0.92). We did not identify statistically significant effects of any covariates on SNP differences in networks #43 and #71.

### Transmission flows analysis

We estimated between and within zone rates of transmission based on transmission pairs identified using TransPhylo (Figure 6, Supplemental Table 5). We found that 68.2% (95% credible interval (CrI): 62.7%, 73.1%) of transmission within the city occurred between, rather than within, zones. Zone 7 in the south-west of the city had the highest percentage of within-zone transmission, followed by Zone 4 in the center of the city. The highest rates of between zone transmission were between Zone 1 and Zone 7 and between Zone 4 and Zone 7. We estimate that 64.5% (95% CrI: 58.9%, 69.9%) of all tuberculosis transmission in Blantyre could be attributed to infectious individuals from Zone 1, Zone 4, or Zone 7. Finally, there was effectively no transmission flow through Zone 3 (posterior mean 1.7 x 10^-5^), a mostly industrial zone in the center of the city with a smaller residential population relative to other zones.

**Figure 6.**
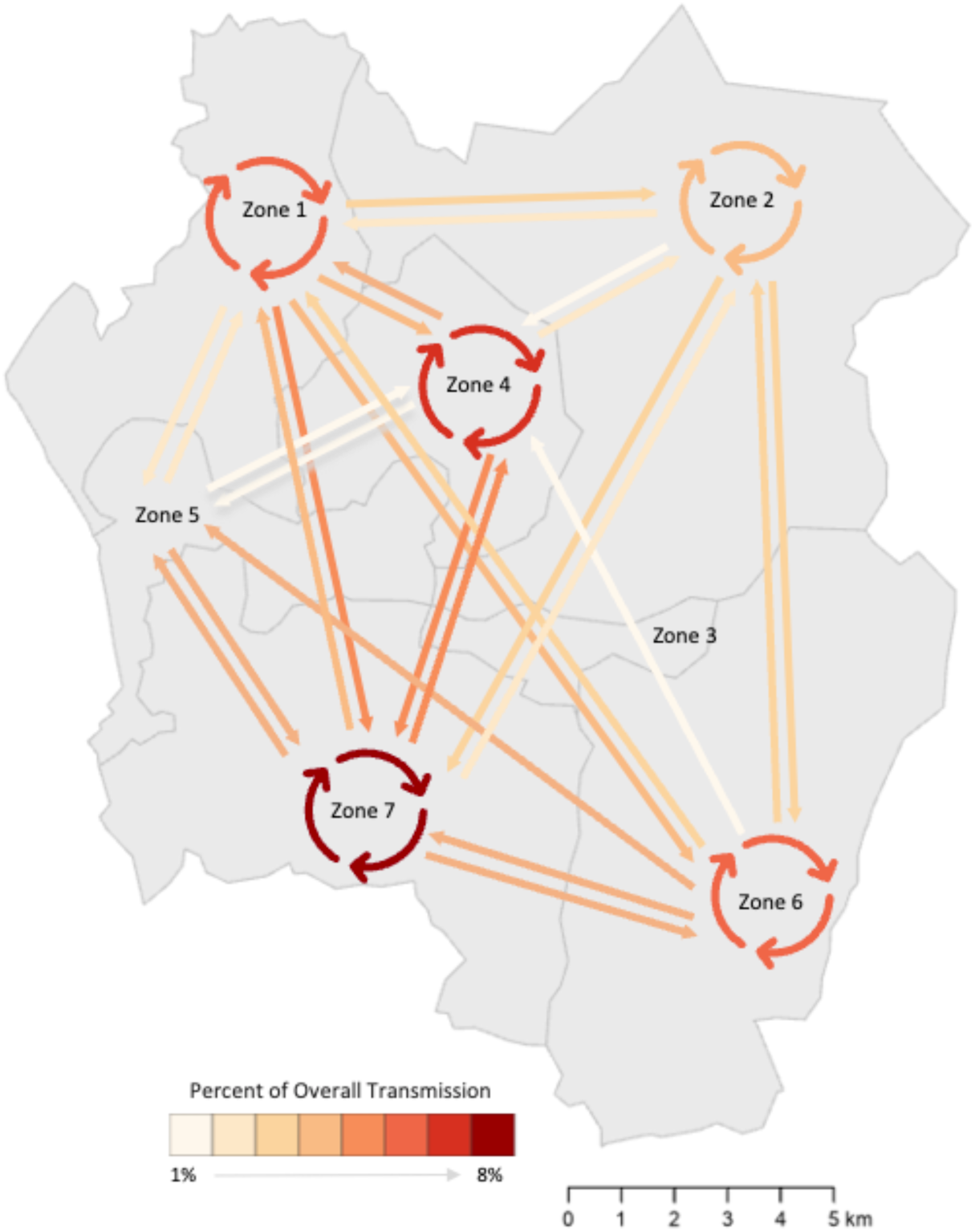
Estimates of the percent of overall transmission occurring within- and between-zones of Blantyre. Arrows point from infector zone to infected zone, and cyclic arrows represent within-zone transmission. Arrows are omitted if there was no evidence of within-zone transmission (e.g. Zone 3) or if there was no evidence of between-zone transmission (e.g. Zone 5 to Zone 3).

## Discussion

This analysis presents novel insights into transmission patterns of *M. tuberculosis* in Blantyre, Malawi between 2015 and 2019, a period when treatment coverage for HIV was being rapidly scaled up and TB notifications were falling. In phylogenetic analyses, 67% of sequenced isolates could be mapped to a putative transmission network. We identified two main characteristics of transmission with implications for the design of public health interventions. First, despite evidence that Blantyre’s tuberculosis epidemic is receding, most detected transmission events occurred between zones, suggesting that disease control efforts must include improved access to tuberculosis services through primary clinics and action to address the social determinants of tuberculosis. Second, we identified a sizeable minority of transmission events that occurred in local outbreaks in distinct areas of the city. Therefore, we anticipate that adding targeted active case finding in these areas is likely to improve tuberculosis control, at least in these neighborhoods.

Geographical location of residence has long been known to be linked to tuberculosis epidemiology, with poor-quality housing, air pollution, undernutrition, crowding, and suboptimal access to healthcare as key drivers of incidence. While the residential proximity of two individuals is an important predictor of membership in the same transmission network (Figure 5A), we found that the majority of transmission events occurred between individuals residing in different zones of the city (Figure 6). High rates of inter-zone transmission are consistent with findings in other high TB/HIV burden settings^(18)^, and indicate that high levels of population mixing play a role in sustaining Blantyre’s tuberculosis epidemic^(45)^. These findings illustrate the continuing need for broad based accessible TB services for all individuals.

These findings also highlight the role that increasingly-available whole genome sequencing data can play in identifying areas of ongoing transmission. Reactive interventions to interrupt transmission may be effective at reducing incidence in these areas, and previous models have suggested that interventions targeted to a single transmission focus could have much broader benefits to reducing incidence in entire city^(6)^. However, the impact of targeted interventions on city-wide tuberculosis incidence depends on the level of connectedness of localized epidemics. In this study, five of the thirteen largest transmission networks were associated with a spatial focus of transmission, though none of the foci overlapped or abutted each other (Figure 4). This finding is distinct from patterns reported from other genomic analyses of tuberculosis in urban settings ^(20, 39)^, and raises questions about the impact of targeted interventions in locations with complex multifocal epidemics.

An important limitation of this study is the relatively low fraction of sequencing (18% of culture-positive isolates over the study period). Diagnostic isolates were available for approximately one-third of eligible cases, and lab contamination further reduced the number of sequenced isolates. This poses a challenge for inferring transmission networks, as most existing methods have been designed for more densely sampled outbreaks. We accounted for this by allowing for a higher number of unsampled intermediate cases in transmission networks, and this introduces additional uncertainty which may have impacted the results of our transmission inference.

In this study, we used genomic and spatial analyses to describe the transmission dynamics of *Mtb* in a high tuberculosis and HIV prevalence city. Our findings reveal the importance of both local and longer range transmission as drivers of tuberculosis transmission in this city. Further modeling is needed to estimate whether targeted screening in areas of focal transmission can be an impactful addition to broader case finding efforts in this city.

## Data availability

The genomic data used in this study will be made available in GenBank upon publication. Additional data used in the analysis (with the exception of patient location and clinic data), will be provided as a file in Supporting Information.

## Ethical Approval

The study protocols were reviewed and approved by the University of Malawi College of Medicine Research and Ethics Committee (#P.12/18/2556), the London School of Hygiene and Tropical Medicine (#16228-4), and Yale (#2000028431).

---

**Supplemental Figure 1:**
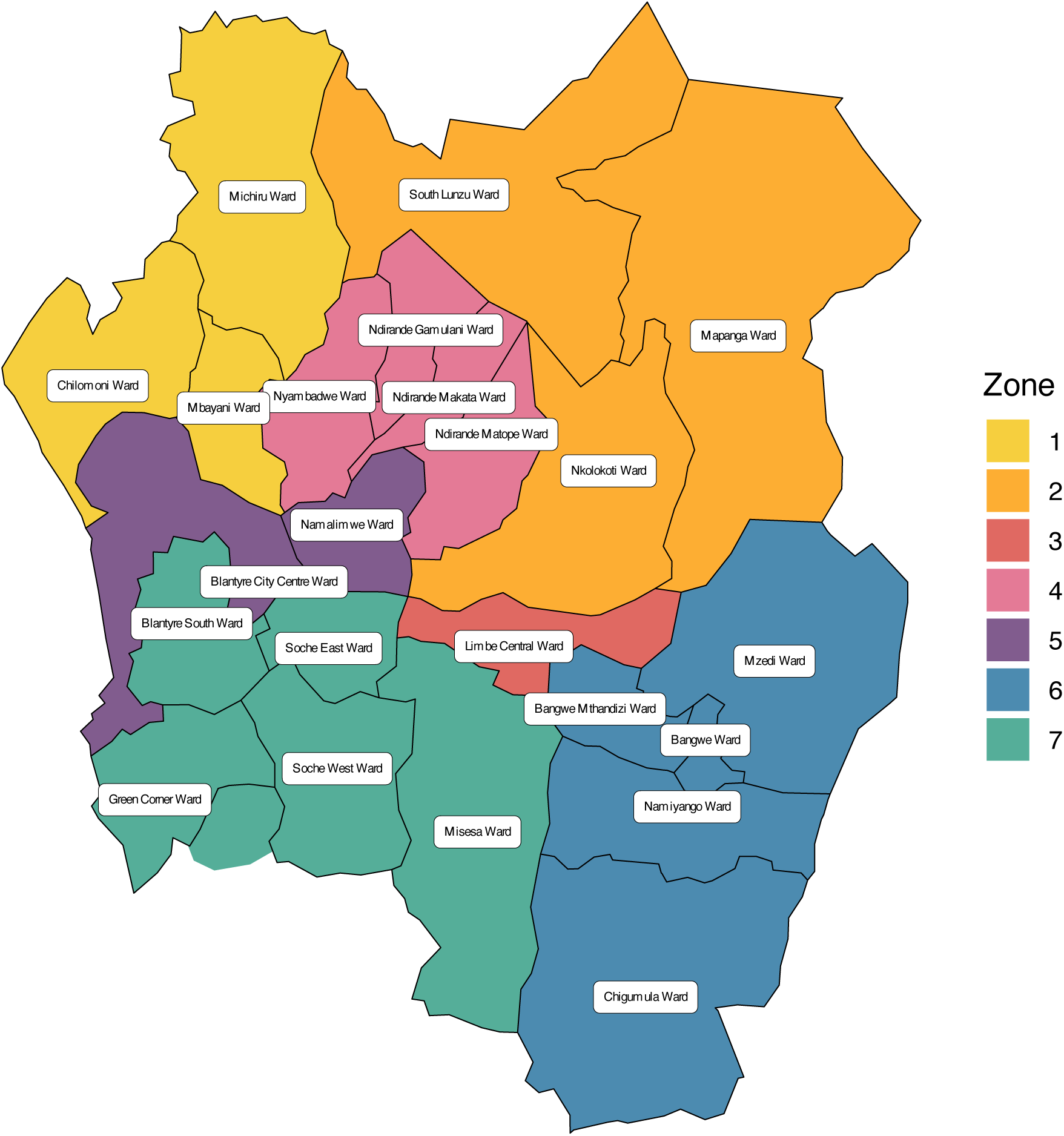
Zones of Blantyre with ward boundaries. Zone 7 has been expanded to include a small area outside the city boundary with a high TB notification rate (unlabeled ward between Green Corner Ward and Soche West Ward).

**Supplemental Figure 2:**
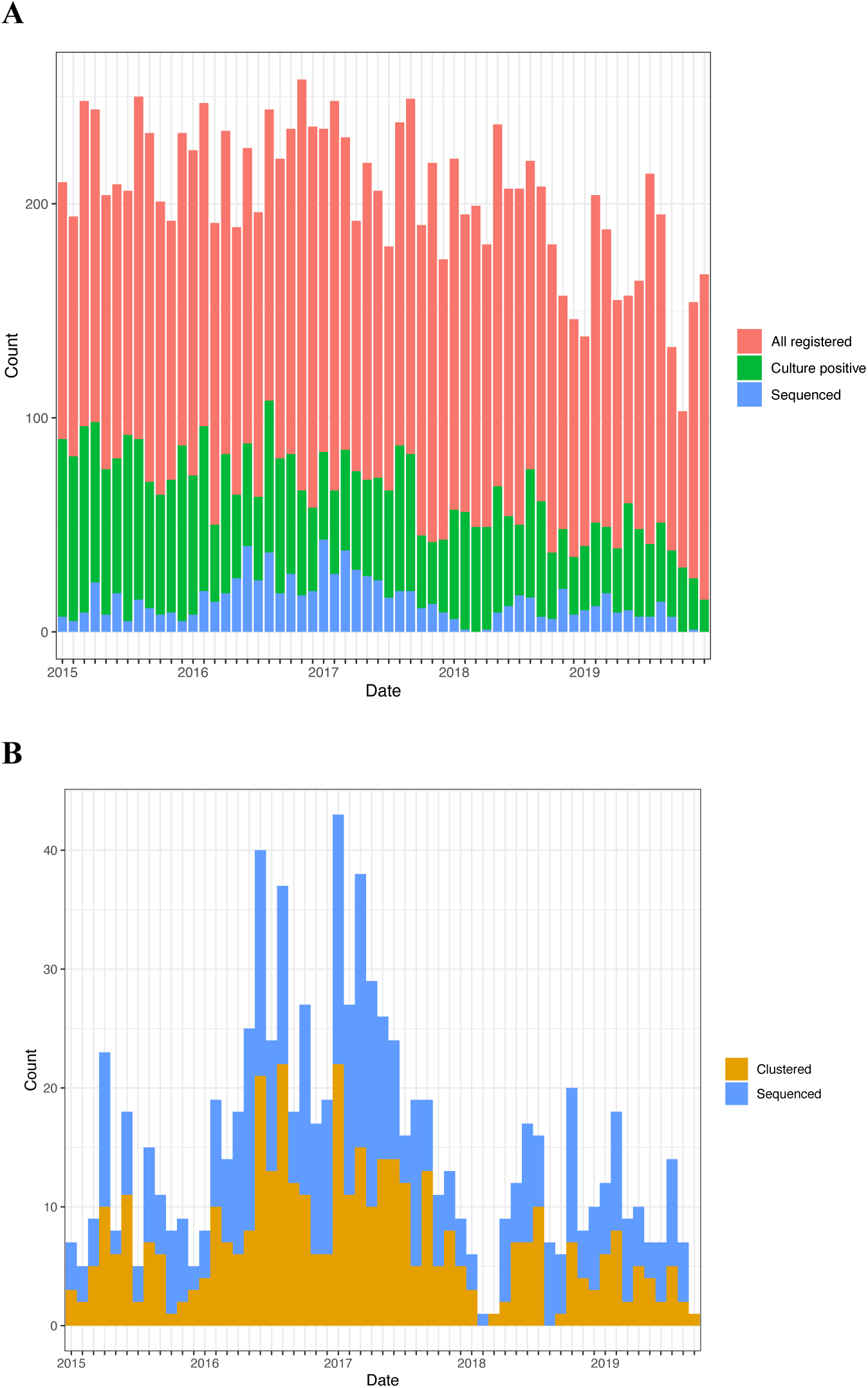
(A) Fraction of cases that were culture-positive and sequenced over time and (B) fraction of sequenced cases belonging to a transmission network (“clustered”) over time.

**Supplemental Figure 3:**
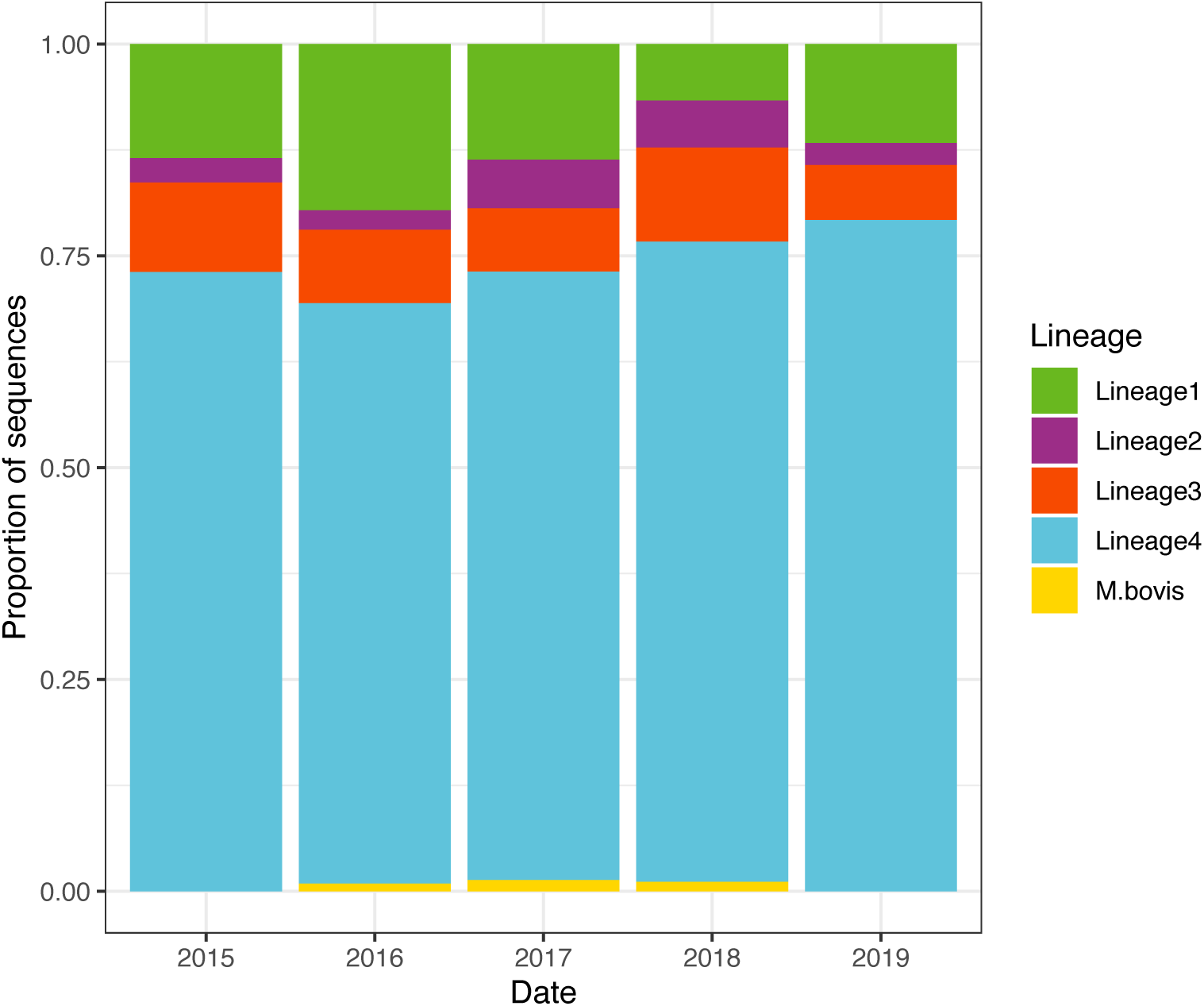
Proportion of sequences belonging to the four major MTBC lineages and *M. bovis*.

**Supplemental Figure 4:**
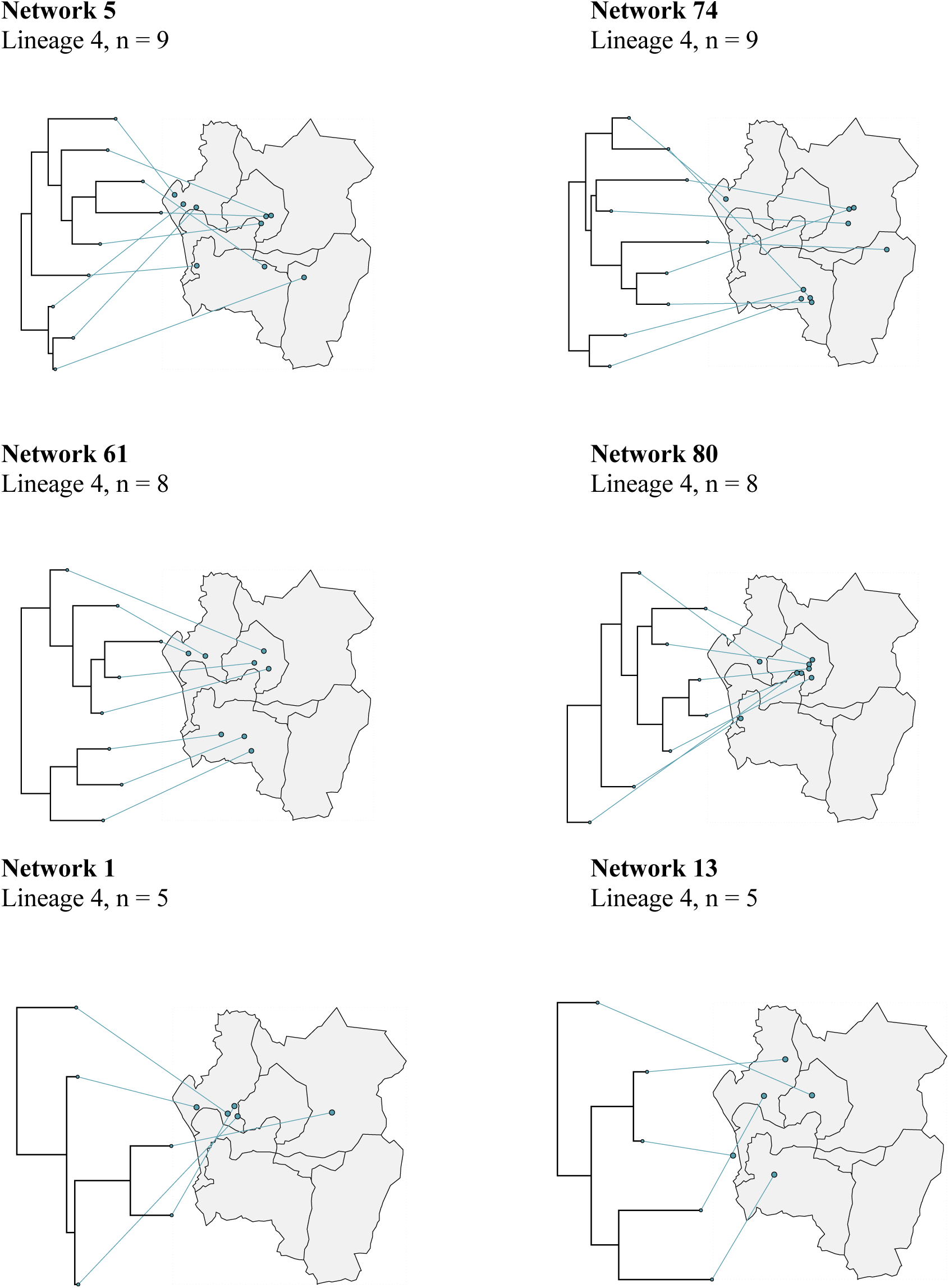
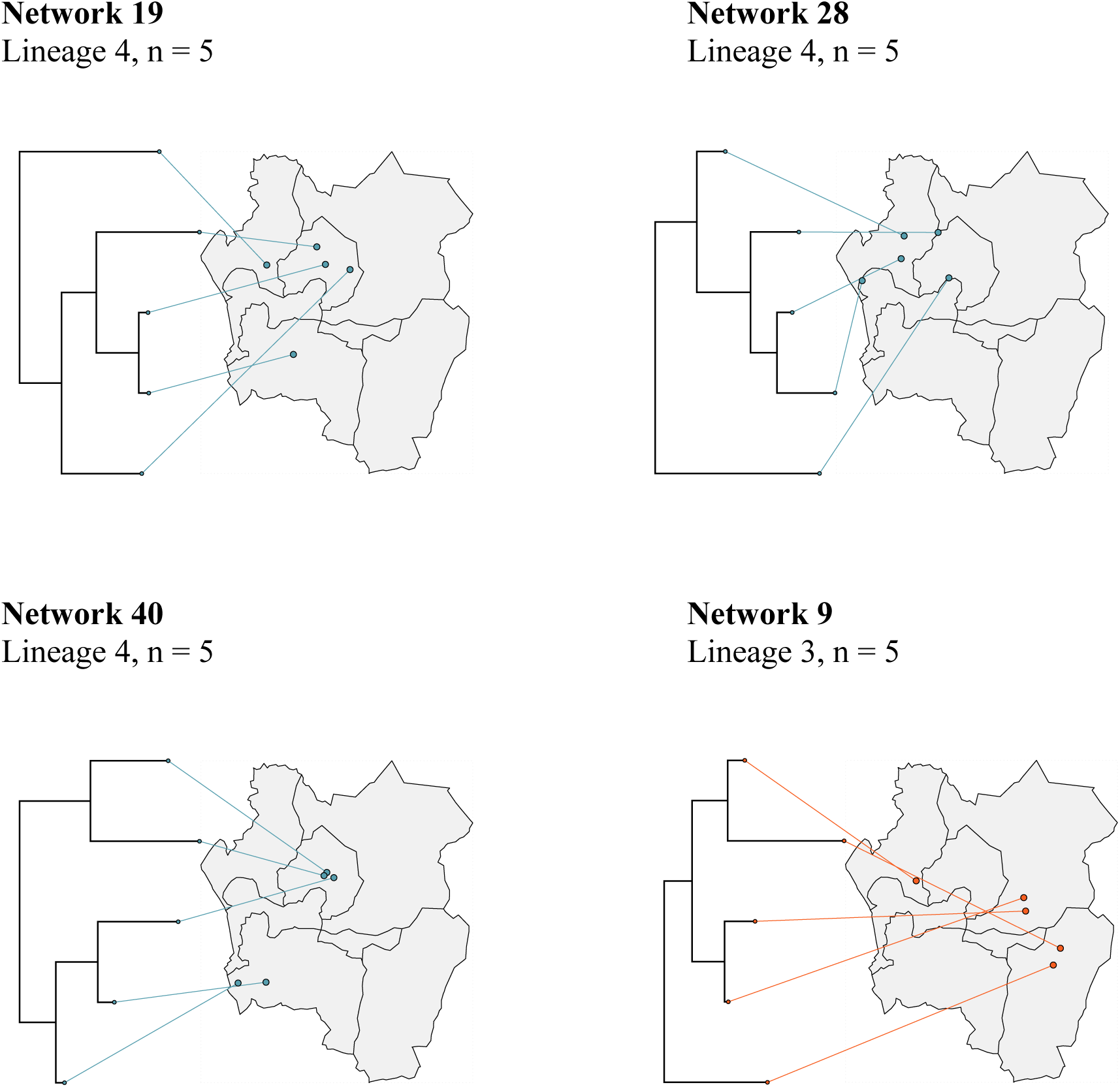
Time-resolved transmission networks with 5-10 sampled cases.

**Supplemental Table 1:**
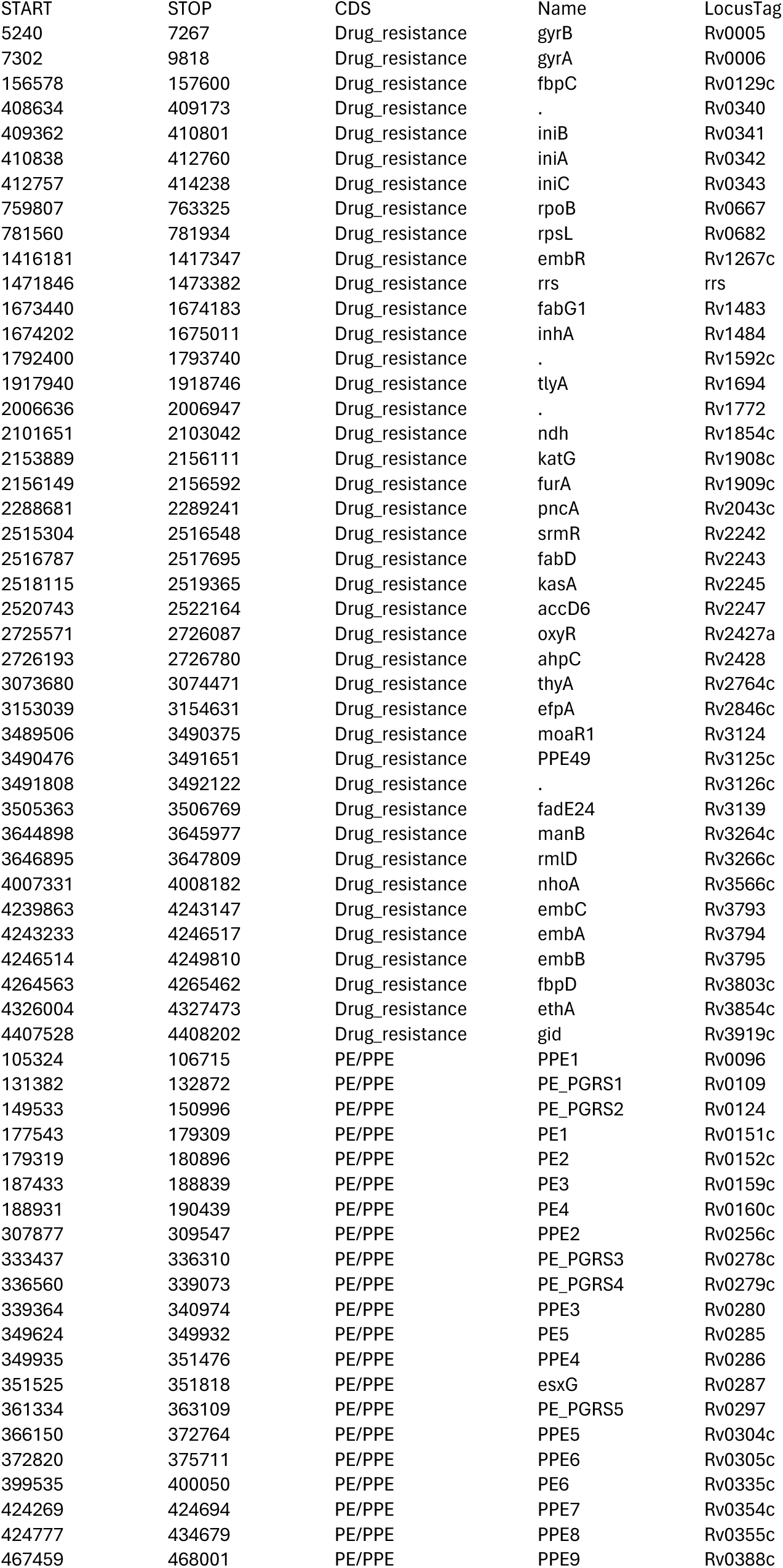
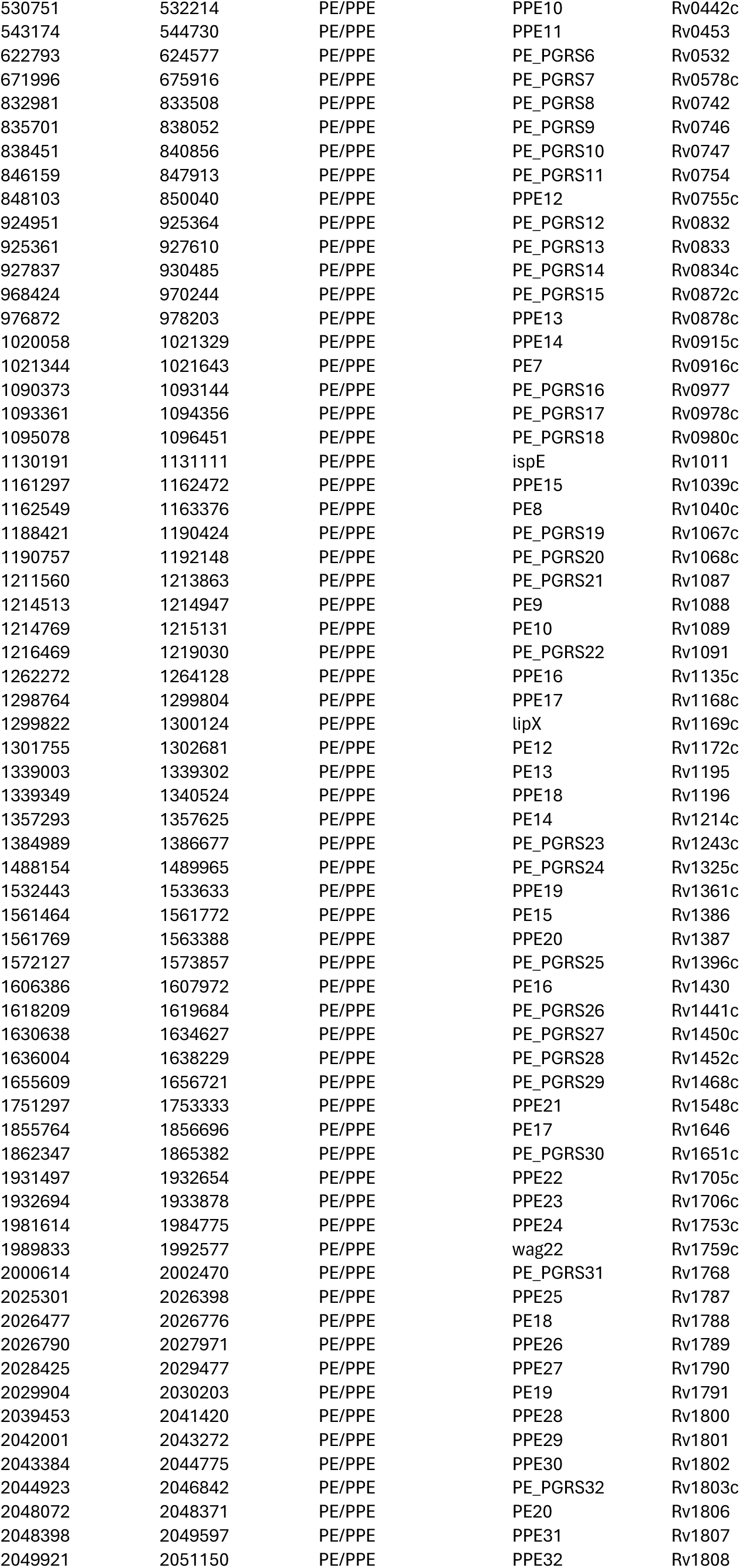
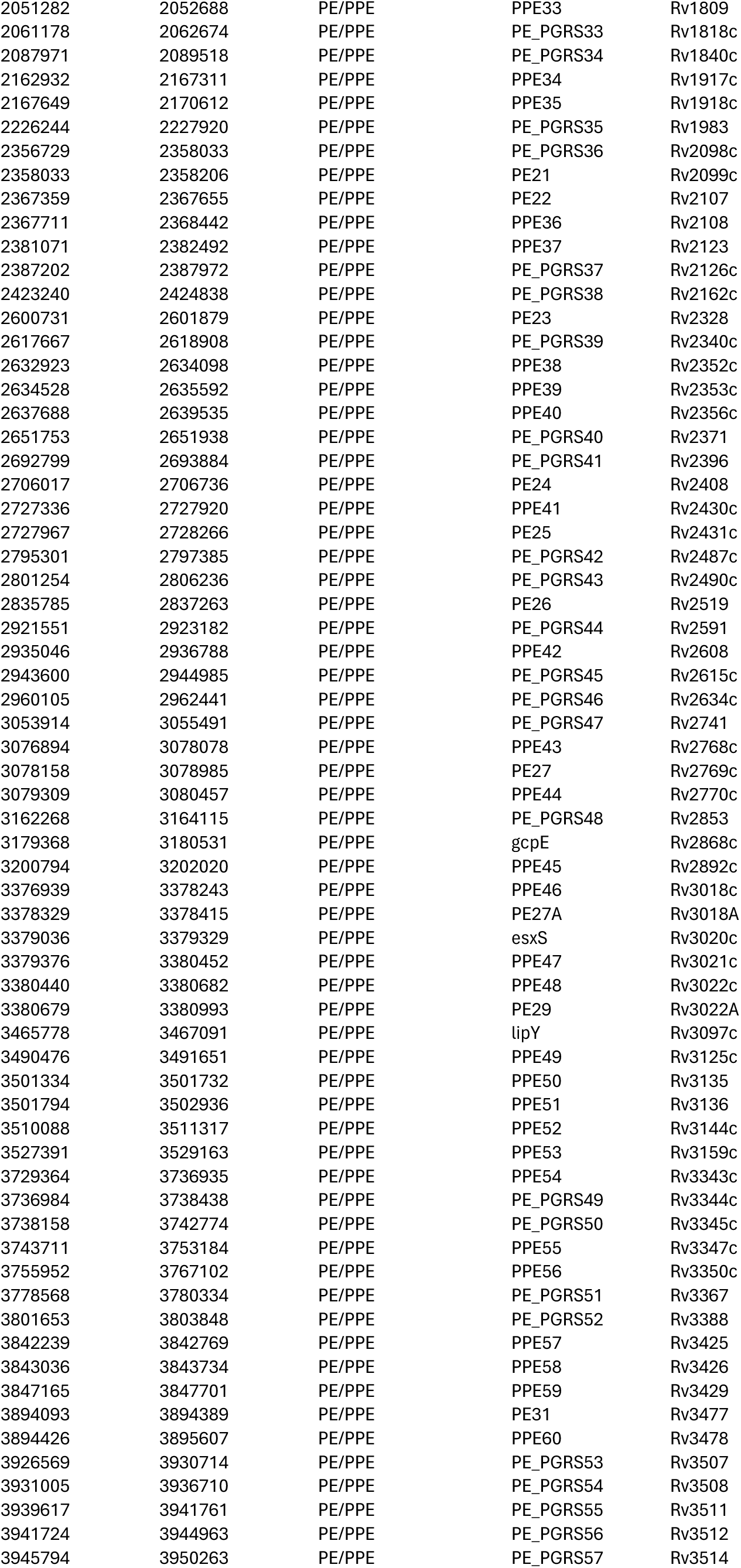
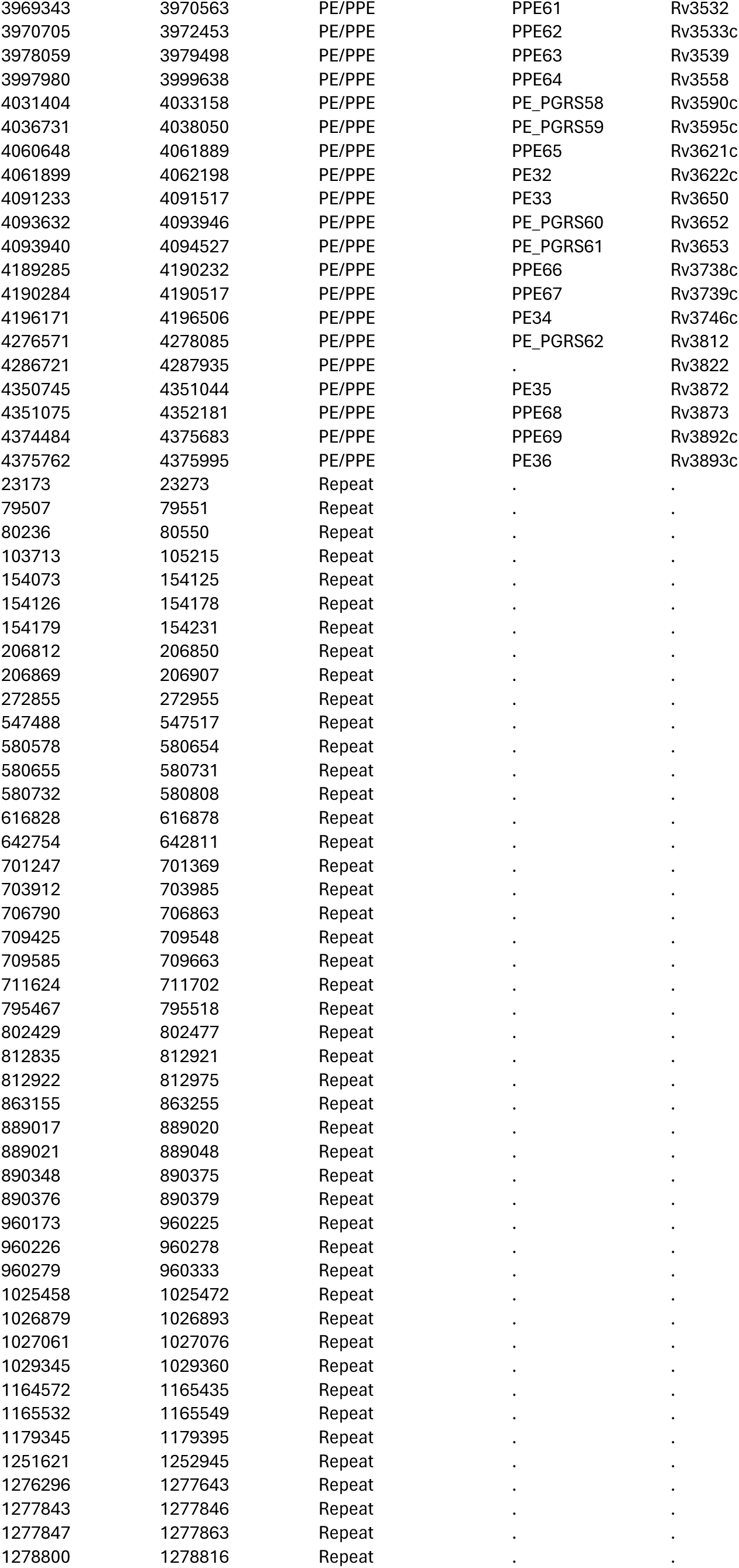
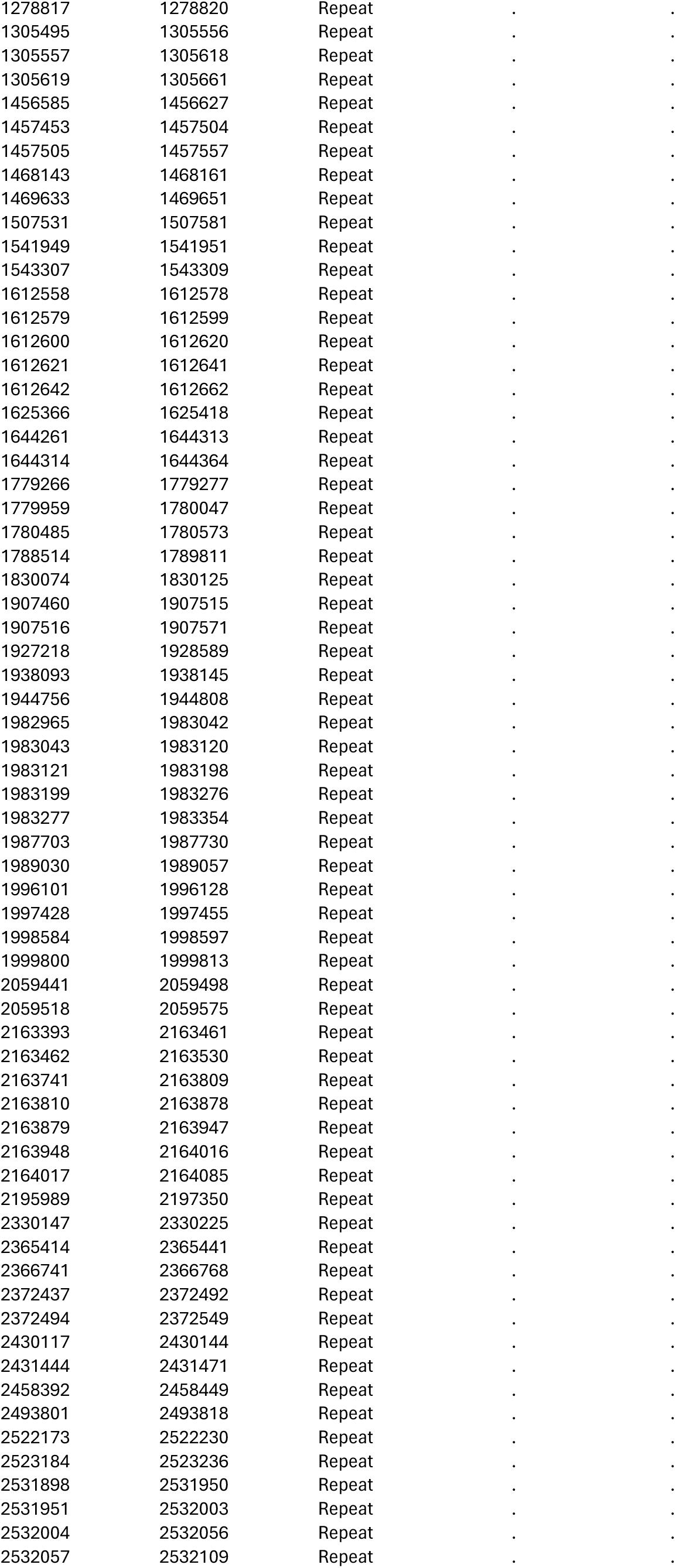
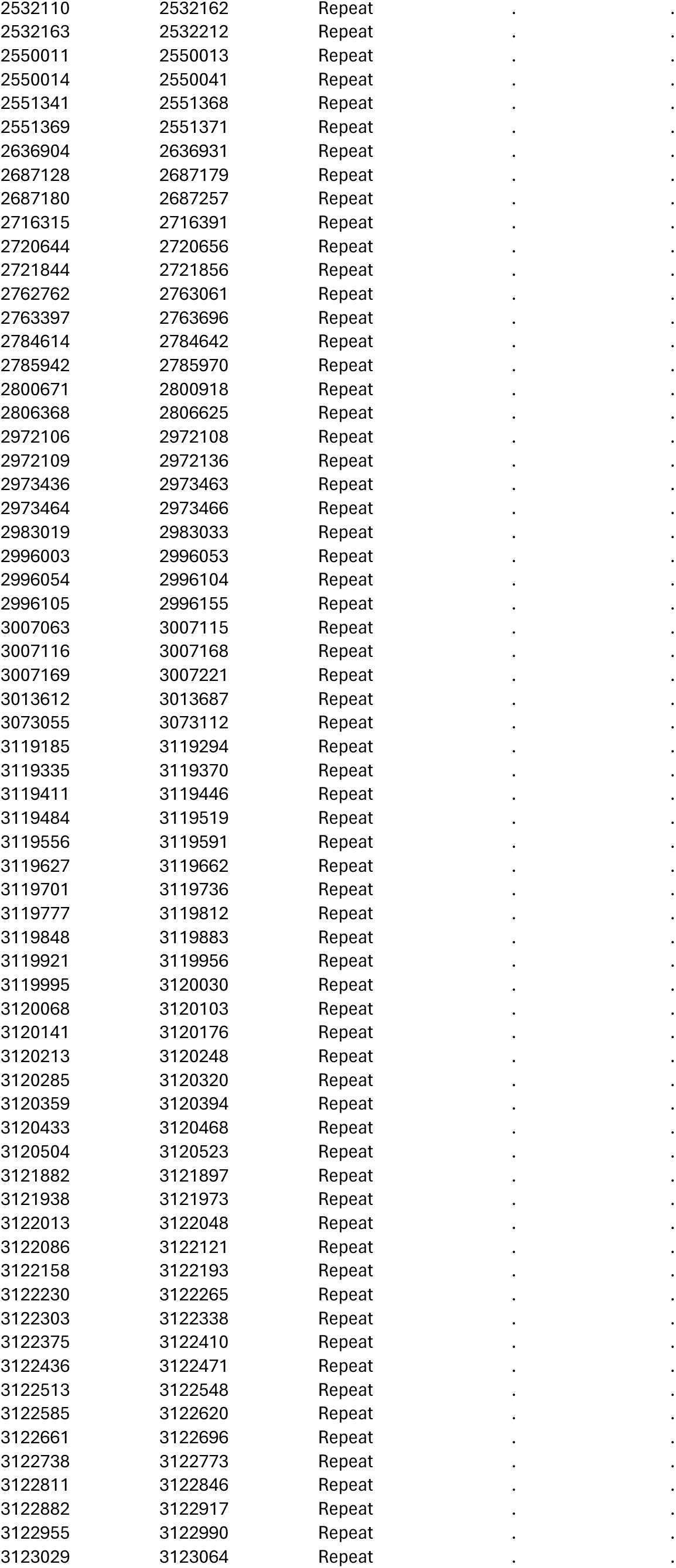
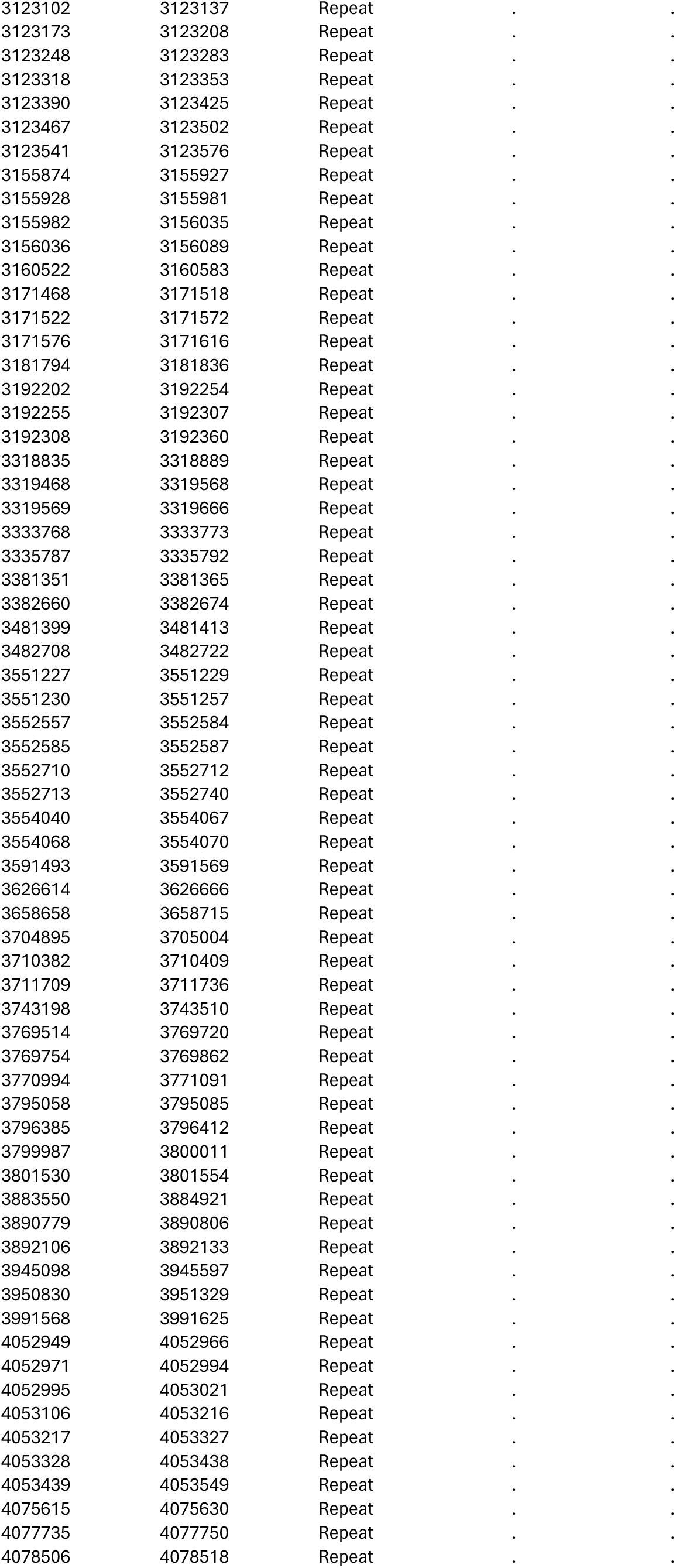
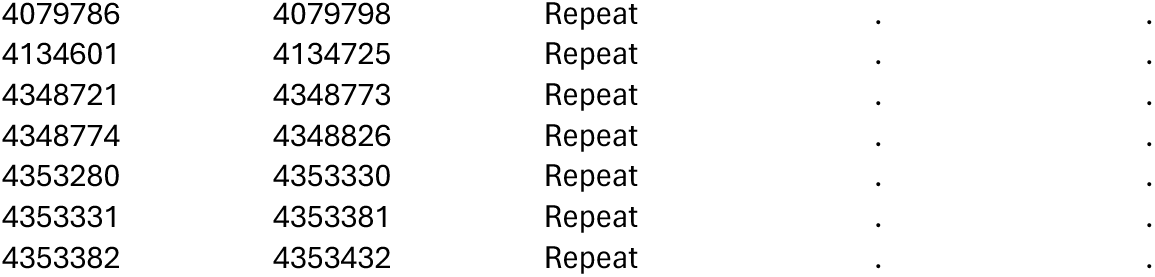
Sites that were excluded from the multi-sequence alignment.

**Supplemental Table 2:** Population size estimates and sequencing coverage by zone.

| Zone | Population (%) | Cases in Zone | Cases with Sequencing Data (%) |
| --- | --- | --- | --- |
| 1 | 181,021 (20.6%) | 1365 | 188 (14%) |
| 2 | 166,388 (19%) | 860 | 88 (10%) |
| 3 | 9,586 (1.1%) | 55 | 2 (4%) |
| 4 | 124,605 (14.2%) | 1249 | 187 (15%) |
| 5 | 42,500 (4.8%) | 305 | 42 (14%) |
| 6 | 144,702 (16.5%) | 1065 | 125 (12%) |
| 7 | 208,753 (23.8%) | 1678 | 209 (13%) |

**Supplemental Table 3:**
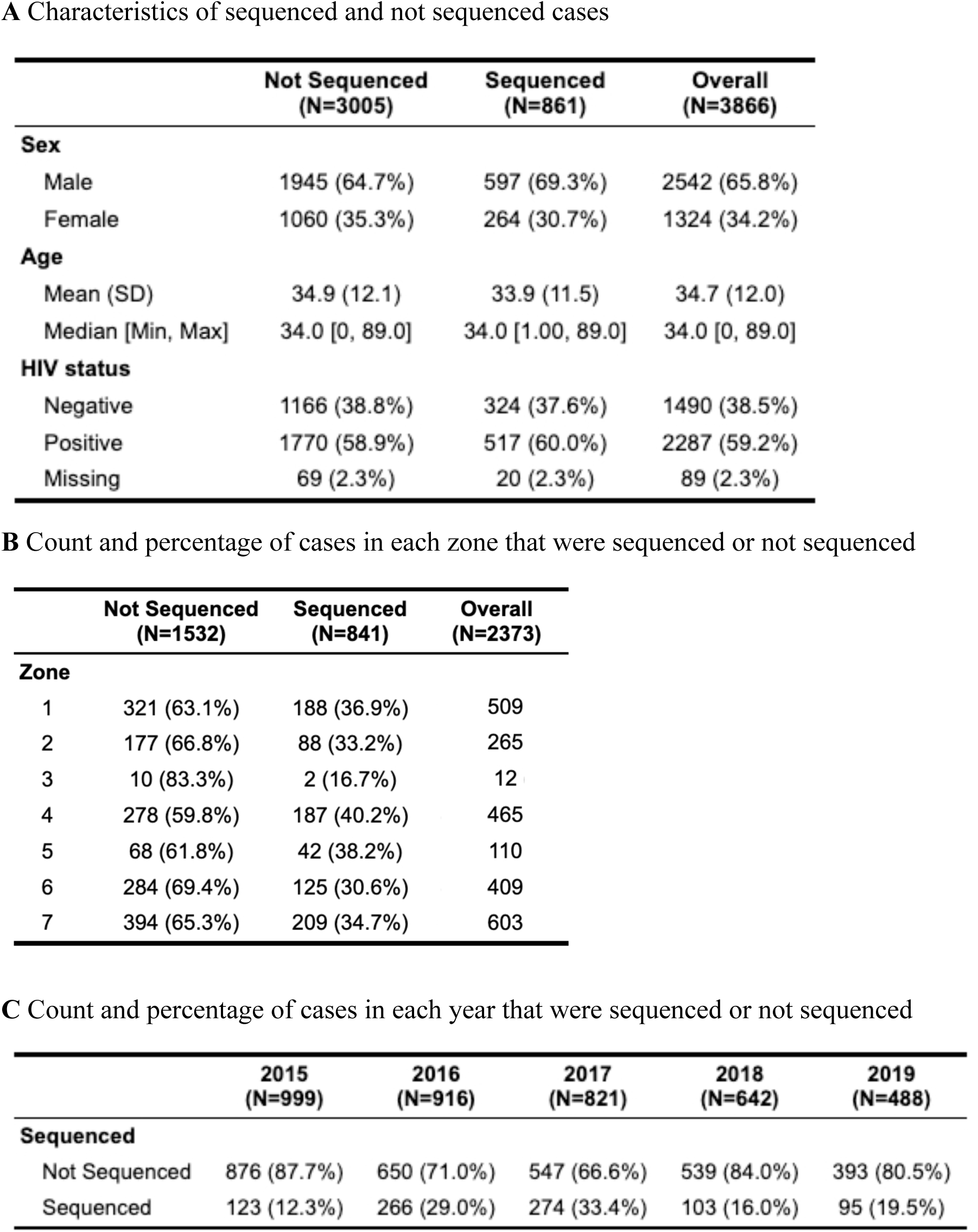
Characteristics of sequenced and not sequenced culture positive TB cases.

|  | **Not Sequenced**<br>**(N=3005)** | **Sequenced**<br>**(N=861)** | **Overall**<br>**(N=3866)** |
| --- | --- | --- | --- |
| **Sex** |  |  |  |
| Male | 1945 (64.7%) | 597 (69.3%) | 2542 (65.8%) |
| Female | 1060 (35.3%) | 264 (30.7%) | 1324 (34.2%) |
| **Age** |  |  |  |
| Mean (SD) | 34.9 (12.1) | 33.9 (11.5) | 34.7 (12.0) |
| Median [Min, Max] | 34.0 [0, 89.0] | 34.0 [1.00, 89.0] | 34.0 [0, 89.0] |
| **HIV status** |  |  |  |
| Negative | 1166 (38.8%) | 324 (37.6%) | 1490 (38.5%) |
| Positive | 1770 (58.9%) | 517 (60.0%) | 2287 (59.2%) |
| Missing | 69 (2.3%) | 20 (2.3%) | 89 (2.3%) |

**B** Count and percentage of cases in each zone that were sequenced or not sequenced
|  | **Not Sequenced**<br>**(N=1532)** | **Sequenced**<br>**(N=841)** | **Overall**<br>**(N=2373)** |
| --- | --- | --- | --- |
| **Zone** |  |  |  |
| 1 | 321 (63.1%) | 188 (36.9%) | 509 |
| 2 | 177 (66.8%) | 88 (33.2%) | 265 |
| 3 | 10 (83.3%) | 2 (16.7%) | 12 |
| 4 | 278 (59.8%) | 187 (40.2%) | 465 |
| 5 | 68 (61.8%) | 42 (38.2%) | 110 |
| 6 | 284 (69.4%) | 125 (30.6%) | 409 |
| 7 | 394 (65.3%) | 209 (34.7%) | 603 |

**C** Count and percentage of cases in each year that were sequenced or not sequenced
|  | **2015**<br>**(N=999)** | **2016**<br>**(N=916)** | **2017**<br>**(N=821)** | **2018**<br>**(N=642)** | **2019**<br>**(N=488)** |
| --- | --- | --- | --- | --- | --- |
| **Sequenced** |  |  |  |  |  |
| Not Sequenced | 876 (87.7%) | 650 (71.0%) | 547 (66.6%) | 539 (84.0%) | 393 (80.5%) |
| Sequenced | 123 (12.3%) | 266 (29.0%) | 274 (33.4%) | 103 (16.0%) | 95 (19.5%) |

**Supplemental Table 4:** Summary of transmission networks used in the DBM analysis.

| Transmission network | Total | HIV positive | Cases within hotspots | Cases within hotspots & HIV positive |
| --- | --- | --- | --- | --- |
| #2 | 25 | 15 (60%) | - | - |
| #43 | 17 | 10 (59%) | 6 (35%) | 4 (67%) |
| #71 | 15 | 3 (20%) | 4 (27%) | 1 (25%) |
| #5 | 9 | 5 (56%) | - | - |
| #74 | 9 | 6 (67%) | 44% | 75% |
| #61 | 8 | 4 (50%) | - | - |
| #80 | 8 | 3 (38%) | 6 (75%) | 2 (33%) |
| #1 | 5 | 3 (60%) | 0 (0%) | 0 (0%) |
| #13 | 5 | 4 (80%) | - | - |
| #19 | 5 | 4 (80%) | - | - |
| #28 | 5 | 1 (20%) | - | - |
| #40 | 5 | 1 (20%) | - | - |
| #9 | 5 | 3 (60%) | - | - |

Transmission networks with spatial foci of transmission are denoted with red text.

**Supplemental Table 5:** Estimated transmission flow (posterior means) between and within zones of Blantyre.

|  |  | Infected Zone |  |  |  |  |  |  |
| --- | --- | --- | --- | --- | --- | --- | --- | --- |
| Infector Zone |  | 1 | 2 | 3 | 4 | 5 | 6 | 7 |
|  | 1 | 0.06 | 0.03 | 0.00 | 0.04 | 0.02 | 0.04 | 0.05 |
|  | 2 | 0.02 | 0.04 | 0.00 | 0.01 | 0.00 | 0.03 | 0.03 |
|  | 3 | 0.00 | 0.00 | 0.00 | 0.00 | 0.00 | 0.00 | 0.00 |
|  | 4 | 0.03 | 0.02 | 0.00 | 0.07 | 0.01 | 0.00 | 0.05 |
|  | 5 | 0.02 | 0.00 | 0.00 | 0.01 | 0.00 | 0.00 | 0.02 |
|  | 6 | 0.03 | 0.03 | 0.00 | 0.01 | 0.01 | 0.06 | 0.03 |
|  | 7 | 0.04 | 0.02 | 0.00 | 0.05 | 0.01 | 0.03 | 0.08 |

